# Effectiveness of a Community-Based Cervical Cancer Stigma Reduction Intervention on Cancer Stigma and Cervical Cancer Screening Uptake in Nepal: A Cluster-Randomized Controlled Trial

**DOI:** 10.64898/2026.09.24.26363856

**Authors:** Priyanka Timsina, Bandana Paneru, Manjari Shrestha, Asika Ghemosu, Seryung Lee, Lisasha Poudel, Yerina S. Ranjit, Donna Spiegelman, Sangini S Sheth, Anne Stangl, Archana Shrestha

## Abstract

Cancer stigma contributes to low cervical cancer screening rates among Nepali women. However, no stigma reduction interventions have been implemented thus far in Nepal. This study assessed the effectiveness of a muticomponent cervical cancer stigma reduction intervention on cancer stigma scores and cervical cancer screening uptake rates among urban Nepali women.

A two-arm, parallel cluster-randomized trial was conducted in Budanilkantha Municipality, Kathmandu District, Nepal. Twelve administrative wards were randomized in a 1:1 ratio to the intervention or control group, with wards serving as the unit of randomization and individual women as the unit of analysis. A total of 310 women aged 30–60 years who had not undergone cervical cancer screening during the previous five years were enrolled. The three-hour intervention included cervical cancer education, a video featuring cervical cancer survivors, participatory group discussions, and myth-versus-fact activities addressing cancer stigma. Cancer stigma was assessed two months after the intervention using the Nepali version of the Cancer Stigma Scale. Cervical cancer screening uptake was assessed six months after the intervention. Data were analyzed using intent-to-treat and as treated approaches with a generalized estimation equation with logistic regression.

From June 2022 to February 2023, 310 women were enrolled. At 2 months, stigma data were available for 248 participants (120 intervention, 128 control). In the intention-to-treat analysis, the odds of cancer stigma were lower in the intervention group than in the control group (OR 0.25, 95% CI 0.08 to 0.8; p=0.022). Odds were also lower for the severity domain (OR 0.31, 95% CI 0.10 to 0.90; p=0.048) and awkwardness domain (OR 0.21, 95% CI 0.08 to 0.56; p=0.001) with no differences in other subdomains. At 6 months, the odds of cervical cancer screening was 3.9 times greater in the intervention group than in the control group (95% CI: 1.1– 13.20, p=0.028). As treated analysis reinforced these findings, with lower odds of stigma (AOR:0.09, 95% CI: 0.01-0.53, p=0.007), severity (AOR:0.16, 95% CI: 0.06-0.42, p<0.001), and awkwardness (AOR:0.1, 95% CI: 0.02-0.44, p=0.02). Cervical cancer screening odds was 4.02 times higher in the intervention group (95% CI:1.1-13.7, p=0.026).

Compared with the control condition, participants allocated to a community-based multicomponent intervention designed to reduce cancer stigma had lower odds of elevated cancer stigma two months after the intervention and higher odds of cervical cancer screening by six months. Larger cluster-randomized trials with more clusters and longer follow-up are needed to assess the reproducibility, durability, and generalizability of these findings.

**Trial Registration:** ClinicalTrials.gov, NCT05489978.

## Introduction

Cervical cancer is the fourth most common cancer in the world, with approximately 604,000 new cases and 342,000 deaths[1]. Nearly 90% of these cases and deaths occur in low- and middle-income countries[1]. Despite being highly preventable through effective measures such as the human papillomavirus (HPV) vaccine, cancer screening, and treatment, cervical cancer remains the second most common cancer among women in Nepal[2]. The country has a crude incidence rate of 16.4 per 100,000 women, with a mortality rate of 11.1 per 100,000 women[2].

The 2010 National Guideline for Cervical Cancer Screening and Prevention in Nepal prioritized screening via the visual inspection acetic acid (VIA) from primary healthcare centers to tertiary levels[3]. Additionally, cervical cancer screening is integrated into the Package of Essential Non-Communicable Diseases (PEN), which was launched in 2017 with an aim to screen, diagnose, treat, and refer to non-communicable diseases at health posts, primary healthcare centers, and district hospitals[4]. Despite sustained governmental efforts to increase screening program accessibility, Nepal reports very low screening coverage (8.2%)[5].

In addition to poor access to these preventive measures and a lack of accurate knowledge [6], cancer stigma is one of the major barriers to screening uptake among women [7–9]. Health-related stigma is a social judgment regarding some health problem, wherein an individual experiences exclusion, rejection, blame, or devaluation[10]. Public stigma involves the social and psychological reactions of society towards a person with some condition that is stigmatized. Self-stigma refers to internalization of the negative beliefs about one’s condition. Stigma by association refers to the reactions that people face when they are associated with a person who has a stigmatized condition. Structural stigma is usually perpetrated within institutions and systems. These forms of stigmatization are connected with one another; public stigma forms the very core of all types of stigma manifestations[11]. Corrigan emphasized public stigma and self-stigma as major obstacles influencing an individual’s decision to seek treatment [12]. Public stigma and self-stigma contribute significantly to suffering, delayed care, higher rates of treatment dropout, reduced program effectiveness, and a diminished quality of life [7,10].

In low-resource settings, stigma is one of the major obstacles to reducing cervical cancer mortality and morbidity [6]. Cancer stigma in Nepal includes beliefs that cancer is fatal, and that cancer can be transmitted through physical contact, fear of social exclusion and various religious and cultural misconceptions[13]. In addition, there is stigma related to HPV with multiple sexual partners which often prevents women from seeking screening services[14]. Mistrust in the healthcare system prevents many from seeking timely care, exacerbating these stigmas and delaying treatment[8]. A recent study in Nepal revealed 23% lower cervical cancer screening coverage among women with higher stigma than among those with low stigma [15].

Existing evidence indicates that most stigma reduction interventions have been developed primarily within the domains of HIV and mental health [16]. There are no stigma reduction interventions implemented in the field of cervical cancer to date. While educational interventions globally aim to increase screening rates [17], those solely addressing knowledge fall short, failing to increase screening coverage [18]. Moreover, existing studies on this topic have predominantly been conducted outside Nepal. Therefore, there is a critical need to develop and evaluate cancer stigma reduction interventions tailored to the Nepali context, assessing their impact on stigma reduction and subsequent improvements in the uptake of cervical cancer screening. This study aims to provide insights for health program managers and stakeholders, aiding in formulating and enhancing strategies to mitigate cancer stigma and promote cervical cancer screening among women in Nepal. The objective of this study was to determine whether a community-based multicomponent stigma reduction intervention reduced cancer and increased cervical cancer screening uptake in Nepal.

## Material and Methods

### Study design

A two-arm, parallel cluster-randomized controlled trial was conducted in Budanilkantha Municipality, Kathmandu District, Bagmati Province, Nepal, between June 2022 and February 2023. Administrative wards were the clusters and the unit of randomization. The allocation ratio was 1:1.

Cluster randomization was used because the intervention involved group-based community activities and discussion of socially shared beliefs and experiences. Randomizing wards rather than individual women was intended to reduce the potential for contamination between intervention and control participants within the same geographic community.

The trial was prospectively registered with ClinicalTrials.gov (NCT05489978). The manuscript was prepared in accordance with CONSORT 2010 guidelines for randomized control trials.

### Study site

The study was conducted in Budanilkantha Municipality, Kathmandu District, Bagmati Province, Nepal. The municipality comprises 13 administrative wards and includes a mixed urban population [19]. Ward 3 was excluded before randomisation because an existing cervical cancer screening programme was operating in that ward. The remaining 12 wards were randomly allocated to six intervention and six control wards.

### Participant recruitment

The inclusion criteria were (a) aged 30-60 years as recommended by National Guideline for Cervical Cancer Screening and Prevention in Nepal for cervical cancer screening 2010[3], (b) married, (c) residents of Budanilkantha Municipality and (d) women who had not undergone cervical cancer screening in the past five years. The exclusion criteria included (a) women with hearing impairment or mental health problems that prevent them from providing written informed consent, (b) pregnant or those less than six weeks postpartum at the time of data collection, (c) women who had resided in Budanilkantha for less than six months due to seasonal work (e.g., in brick kilns) or as visitors to the families of Budanilkantha residents, (d) women already diagnosed with cervical precancer, and (e) prior cancer affected women leading to hysterectomy.

Female community health volunteers (FCHVs) and elected female ward representatives were oriented to the study objectives, eligibility criteria, recruitment procedures and their roles in community mobilisation. They disseminated study information and helped identify women who expressed interest. Eligibility was subsequently assessed by study staff.

### Randomization, allocation, concealment and blinding

The 12 eligible wards were randomly allocated in a 1:1 ratio to either the cervical cancer stigma-reduction intervention or the control group using a computer-generated random sequence. The randomization sequence was generated using Stata version 14 by an investigator (AS) who was not involved in implementation, participant enrolment, outcome assessment, or data analysis.

The generated sequence was then emailed to another investigator (PT), who assigned the wards to the intervention or control group according to their serial numbers in the randomization sequence. Thus, the ward was the unit of randomization, and all eligible women within a ward received the intervention or control condition corresponding to their ward’s allocation.

### Intervention

The women in the intervention group received one day 3-hour sessions with 12 participants, facilitated by trained public health professionals with two years of experience in health education interventions. The intervention comprised two broad components.

#### Cervical cancer education

Facilitators conducted a 20-minute PowerPoint slide-aided presentation on cervical cancer, including signs, symptoms, preventive measures, and available treatment services. The slides predominantly featured images with minimal text, considering potential literacy challenges within the community.

#### Stigma reduction

Facilitators addressed cancer stigma using three step-wise sessions. First, a 20-minute video featuring cervical cancer survivors from both urban and rural settings was shown, where the survivor narrates her life experiences. The video emphasized the importance of seeking medical help when unusual symptoms appear, encouraging individuals not to feel embarrassed and to be screened and treated early. The protagonist in the video reflects the women’s socio-demographic characteristics intended to generate parasocial relationships with the viewers. Parasocial relationship refers to a one-sided relationship where individuals feel connected with others despite having no direct interaction [20]. When individuals identify with the characters in the media and relate to them, it helps them connect the information to their own personal life experiences[21].This aims to change attitudes and behaviors related to the severity, avoidance, and personal responsibility domains of cancer stigma. Second, the facilitators organized a group of four participants for a two-hour discussion. During this session, they shared personal experiences related to barriers in cervical cancer screening and explored the drivers, facilitators, types, and consequences of perceived or community stigma. The group documented their discussion on chart paper, and the leader presented the findings to the larger group. Third, the facilitator conducted 20-minute myths, misconceptions, and facts to address severity, financial discrimination, and policy opposition domain using flashcards.

The intervention was guided by the Stigma Mechanisms in Health Disparities Framework, which describes how public stigma accounts for disparate outcomes among stigmatized and non-stigmatized people[22]. The intervention arm was invited to participate in the cervical cancer screening services by informing about the date and place of the screening via telephone. Among the 156 participants in the intervention group, 18 did not receive our calls, 5 phones were switched off, 6 were unreachable, and 2 were wrong numbers. A detailed intervention matrix is given in Table 1.

**Table 1.** Intervention Matrix.

| Objectives | Contents | Time | Media | Target |
| --- | --- | --- | --- | --- |
| 1. Change in knowledge |  |  |  |  |
| To make women aware about the increasing burden of cervical cancer, its prevention and treatment measures. | Awareness raising programs on cervical cancer and cervical cancer screening<br>Cervical cancer burden in Nepal, signs and symptoms, preventive measures available, lifestyle modifications, HPV vaccination, early diagnosis, screening and treatment.<br>Screening measures available in Nepal, accessibility and availability of screening services, interval of screening, financial determinant, treatment services available in Nepal | 20 min | Presentation slide with projector | Knowledge and awareness on cervical cancer and screening |
| 2. Change in stigma: To reduce individual, interpersonal and sociocultural stigma on cervical cancer among women improve well-being. |  |  |  |  |
| To change individual level internalized stigma, we shall show them a video of a cervical cancer survivor and narrate live experiences of cervical cancer survivors. | Video of a person facing cervical cancer what symptoms she got, how she dealt with it and how she overcame it. (Highlights on importance of screening if any unusual symptoms are found and not to be embarrassed and seek medical help and treatment as early as possible) | 20 min | Parasocial relationship | Target: Severity, avoidance, personal responsibility |
| To enhance social network, social interaction and social support we shall participatory learning techniques | Personal stories: Share stories of women about the barriers of going to screening programs.<br>Voluntary self-reflection of any stigma they perceive on cervical cancer, or their community perceives on cervical cancer. | 2 hours | Participatory discussion | Target: Avoidance, Awkwardness |
|  | Group Discussion: Form a group of 4 and discuss the drivers of stigma, facilitators of sigma, types of stigmas prevalent in your community, consequences of stigma and present it by themselves. |  |  |  |
| To correct myths and misconceptions and to challenge negative perceptions based on 6 domains of cancer stigma scale. | Myth: Cervical cancer is fatal<br>Fact: Cervical cancer can be cured if detected and treated at early stage.<br>Myth: Cervical cancer is contagious<br>Fact: Cervical cancer itself is not contagious. HPV infection is sexually transmitted and persistent infection with oncogenic HPV types can cause cervical cancer.<br>Myth: I don't need to get screened because I don't have any symptoms<br>Fact: I need to get screened in every 5 years irrespective of my symptoms.<br>Myth: HPV isn't that common it affects people with multiple sexual partners.<br>Fact: Approximately 80% of men and women are infected at some point in their lifetime.<br>Myth: I don't want to get screened because if I have cervical cancer, it cannot be treated.<br>Fact: Screening tests help prevent cervical cancer screening by detecting subnormal cells in the cervix. So, women who don't get screened might miss the opportunities to detect abnormal cells early when treatment is very effective. | 20 mins | Myth vs Facts cards | Target: Severity, financial discrimination and policy opposition. |

### Control group

Participants in the control group received usual care consisting of information about the availability, date, and location of cervical cancer screening services. They did not receive the additional stigma-reduction components provided to the intervention group, including structured cervical cancer education, the cervical cancer survivor video, participatory group discussion, and myth-versus-fact activities. The control condition therefore represented the usual screening-information approach against which the multicomponent intervention was evaluated.

### Data collection tool and variable measurement

#### Baseline Assessment

Baseline data were collected through face-to-face interviews using a structured electronic questionnaire administered on tablets through REDCap. The baseline questionnaire captured sociodemographic characteristics, reproductive and obstetric characteristics and cancer stigma. Sociodemographic variables included age (years), ethnicity (Janjati, Brahmin/Chhetri, or other), educational attainment (no formal education or ≥1 year of formal education), occupation (homemaker, salaried employment, agriculture, daily wage labour, self-employment, or other), husband’s occupation (foreign employment, salaried employment, agriculture, daily wage labour, self-employment, or other) and annual household income (US$). Reproductive and obstetric variables included age at marriage, age at menarche, age at first pregnancy, menopausal status, age at menopause, number of children, number of pregnancies, history of caesarean section and shortest birth interval (years).

#### Follow-up Assessment

Endline assessment was conducted two months after completion of the intervention. At this assessment, cancer stigma was remeasured using the same instrument and administration procedures as at baseline. Cervical cancer screening uptake was assessed six months after the intervention.

### Outcomes

#### Cancer stigma

Cancer stigma was assessed using the Nepali version of the Cancer Stigma Scale (CASS), a validated tool designed for this population.[23]. Cancer stigma was re-measured in both groups two months after the intervention. The two-month time frame was chosen on the basis of studies indicating that stigma reduction interventions have weaker effects over prolonged periods [18]. The CASS comprises 25 six subdomains of cancer stigma: severity domain assesses the perceived consequences of a cancer diagnosis and the likelihood of recovery, personal responsibility domain assesses how much a person’s actions are believed to contribute to their cancer, awkwardness domain assesses the comfort levels of cancer patients, avoidance domain assesses the extent to which people avoid cancer patients and maintain physical distance, the policy opposition domain assesses the perception of government and public responsibility for cancer patients’ treatment and care, financial discrimination domain assesses the extent to which cancer patients are expected to benefit from financial services such as banks and insurance. The participants responded on a 6-point Likert scale (1 = strongly disagree to 6 = agree). The mean scores for each domain were calculated after reversing the scores of the five positive features of the policy opposition and awkwardness domains. Higher scores correspond to higher stigma levels [24]. Subsequently, the mean score was dichotomized into having stigma (>=3) or no stigma (<3).

#### Cervical cancer screening uptake

Cervical cancer screening uptake was assessed six months after the intervention and was defined as having undergone cervical cancer screening during the six-month follow-up period. Screening status was ascertained using a two-stage approach. First, participants’ screening records were verified against administrative records from cervical cancer screening camps conducted in Budanilkantha Municipality. For participants for whom no screening record was identified in the available municipal screening databases, follow-up was conducted by telephone to determine whether they had undergone cervical cancer screening at another health facility or screening site during the follow-up period.

Participants were classified as having screened if screening was documented in the available administrative records or was reported by the participant during telephone follow-up.

Participants for whom no evidence of screening was identified through either source were classified as not screened.

### Sample size

The target sample size was 310 participants (155 per arm), calculated to detect a 10% decrease in mean stigma score between groups after the intervention. The calculation used a mean score of 2.6 and SD 0.6 from a previous study [17], a two-sided alpha of 0.05, 85% power, a design effect of 1.24 based on an intracluster correlation coefficient of 0.01 and an average cluster size of 25, and 30% anticipated loss to follow-up [25]. The primary analysis subsequently used the prespecified CASS threshold of ≥3 to define elevated stigma; this distinction between the scale used for sample-size planning and the binary analysis is considered when interpreting the findings.

### Statistical analysis

Data were analyzed using Stata/MP version 14.0 (StataCorp, College Station, TX, USA). The sociodemographic and reproductive health characteristics of the intervention and control groups were summarized using frequencies (percentages) for categorical variables and means (standard deviations) for continuous variables.

The primary analysis followed the intention-to-treat (ITT) principle. Participants were analyzed according to the intervention group to which they were randomized, regardless of whether they received or completed the intervention. Participants who Did not receive intervention were included in the primary analysis within the respective treatment group they have been assigned to at randomization (“as randomized”). For the 2-month cancer stigma outcome, 248 participants contributed outcome data (120 intervention and 128 control). For the 6-month cervical cancer screening outcome, 244 participants contributed outcome data (125 intervention and 119 control). Participants without the corresponding outcome measurement were excluded from that specific outcome analysis. The extent and pattern of missing outcome data were considered when interpreting the findings.

A secondary as-treated analysis was conducted as per the intervention. Twenty-three participants who had been randomized to the intervention group but declined the intervention were classified with the control group for this analysis. For the cancer stigma outcome, 248 participants were included in the as-treated analysis, comprising 105 participants classified in the intervention group and 143 in the control group. For the screening outcome, 244 participants were included, comprising 101 participants classified in the intervention group and 143 in the control group. As randomization was violated, adjustments were made for confounding variables in this analysis, including age, level of education completed, religion, ethnicity, work status, husband’s work status, husband living together, personal income, family income, history of menopause, number of pregnancies and age at first pregnancy in as- treated analysis. As this is a c-RCT data would be clustered within wards we assume to have data clustered within a ward because of similar sociodemographic and cultural characteristics. Thus, the outcomes among individuals in the same ward would be correlated. To address this, generalized estimating equations (GEE) with a binomial distribution and logit link function, exchangeable working correlation and the robust variance was used to examine the associations between intervention status (yes/no) and cancer stigma (mean cancer stigma score <3 and ≥3) and screening uptake (yes/no)[26]. GEE corrects for correlation of responses for participants clustered within a ward, and robust variance estimators provide assurance for valid inference under possibly mis-specified correlation structure. Odds ratios (ORs) with 95% confidence intervals (CIs) and p-values were reported. All p-values less than 0.05 were considered statistically significant.

### Ethical Consideration

The study was conducted in accordance with the CONSORT 2010 guidelines for randomized control trials. Ethical approval for the study was obtained from the Institutional Review Committee (IRC) of Kathmandu University School of Medical Sciences (IRC reference number: 42/22). The study was registered at ClinicalTrials.gov (Identification Number: NCT05489978). Written informed consent was obtained from literate participants who were able to read and write. For participants who were illiterate, verbal informed consent was obtained, and the consent form was signed by a witness of the participant’s choice. Throughout the study, ethical principles were strictly upheld, with particular emphasis on confidentiality, voluntary participation, and participants’ right to withdraw from the study at any time without any consequences.

## Results

### Participants Flow

Between June 2022 and February 2023, a total of 310 eligible women were enrolled from 12 randomised wards. Six wards were allocated to the intervention group and six to the control group, with 156 women enrolled in the intervention wards and 154 in the control wards at baseline. Follow-up data for the stigma outcome were available for 248 participants, comprising 120 participants in the intervention group and 128 in the control group. Loss to follow-up was attributable to difficulties identifying participants due to discrepancies between commonly used names and citizenship names, incorrect telephone numbers, unanswered or switched-off telephones, competing commitments and withdrawal from further participation.

For the screening uptake outcome, follow-up data were available for 244 participants, including 101 in the intervention group and 143 in the control group. Reasons for loss to follow-up included failure to receive the study call, switched-off telephones, incorrect telephone numbers, and participants being unreachable. The flow of participants through enrolment, allocation, follow-up and analysis is presented in the CONSORT flow diagram (Fig 1).

**Fig 1.**
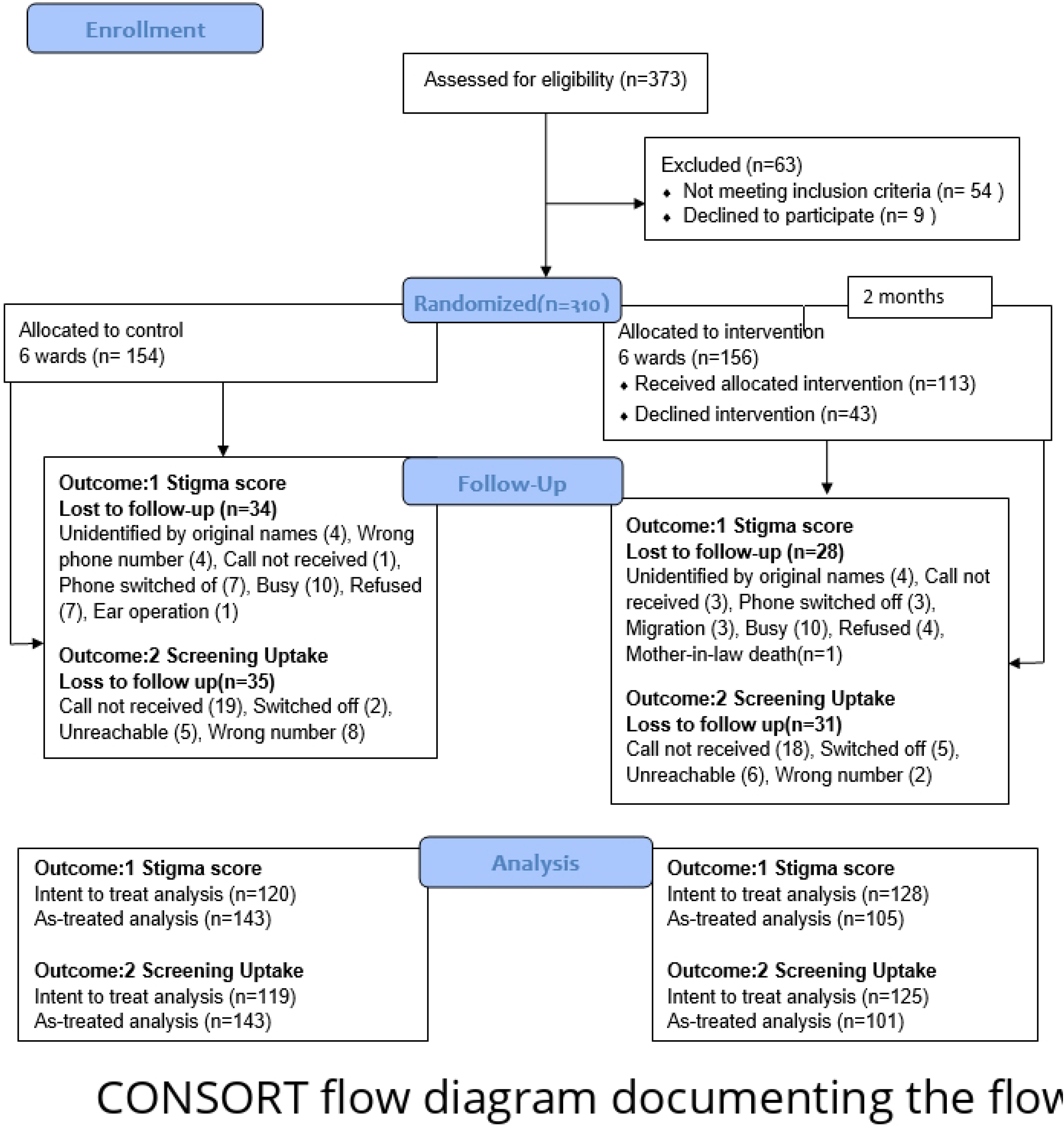
CONSORT flow diagram documenting the flow of participants through enrolment, allocation, follow-up and analysis.

### Sociodemographic and reproductive characteristics of study participants

The mean age of the participants was 41.5±7.8 years. More than a quarter of the participants had no formal education (28%). There were no differences between the groups in sociodemographic characteristics including age, ethnicity, educational status, personal income, family income, or reproductive health characteristics such as age at first pregnancy, number of pregnancies or history of menopause between the intervention and control groups at baseline. The control group had a greater percentage of Hindus (85%) than did the intervention group (76%). Table 2 presents the baseline characteristics of the study participants.

**Table 2.**
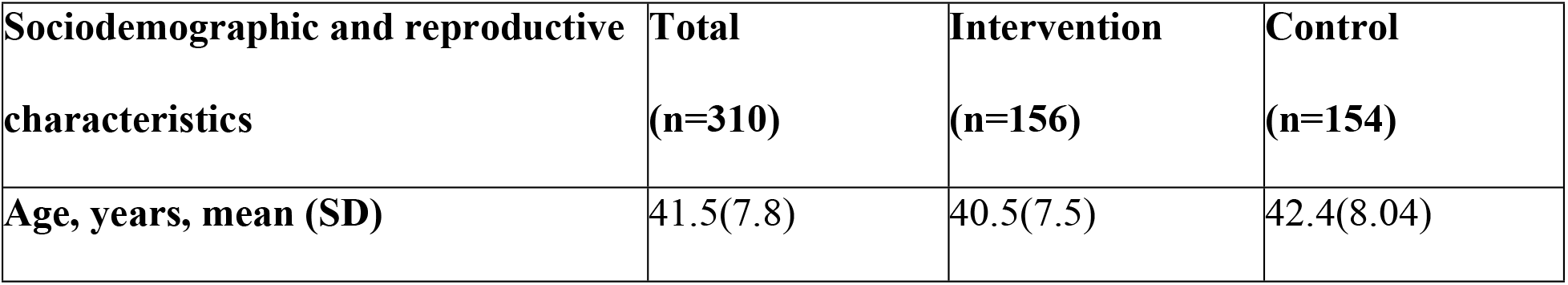
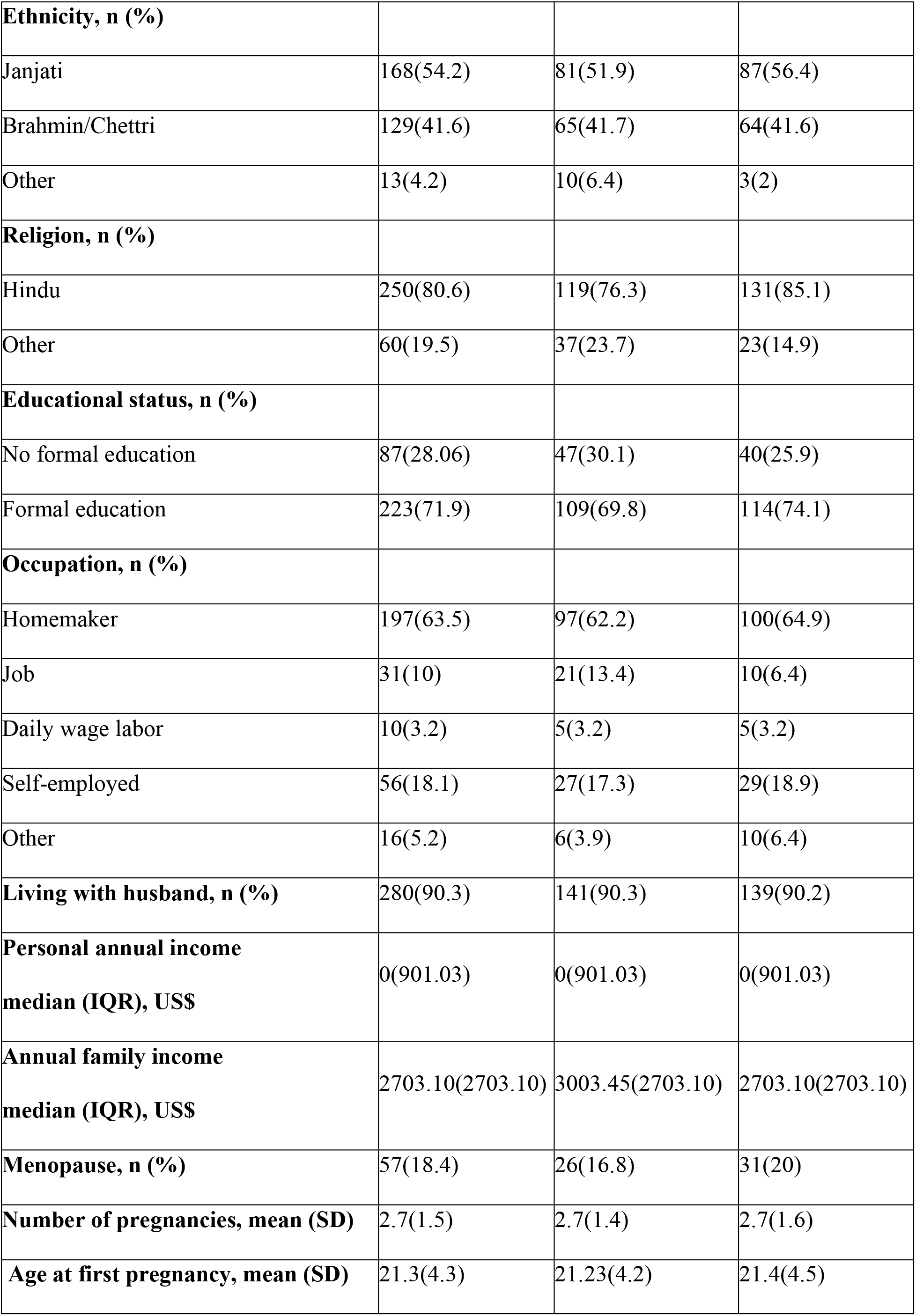
Sociodemographic and reproductive characteristics of study the participants by group.

### Cancer stigma score

Overall, the participants had a low mean Cancer Stigma Scale (CASS) score, with considerable variation observed across all six subdomains (Table 3). At baseline, the mean CASS score was 2.7±0.7 for the intervention group and 2.6±0.7 for the control group. Both groups showed a reduction in CASS score at follow-up, with the intervention group having a lower score mean 1.4±0.6) than the control group (1.9±0.8).

**Table 3.** Mean cancer stigma score in each domain in the intervention and control groups at baseline and 2-month follow-up.

| **Cancer stigma domains** | **Baseline (n=310)** |  |  | **Follow-up (n=248)** |  |  |
| --- | --- | --- | --- | --- | --- | --- |
|  | **Intervention,**<br>**mean (SD)** | **Control,**<br>**mean (SD)** | **P-**<br>**value** | **Intervention,**<br>**mean (SD)** | **Control,**<br>**mean (SD)** | **P-**<br>**value** |
| **Awkwardness** | 2.3(1.4) | 2.1(1.4) | 0.492 | 1.2(0.8) | 1.7(1.3) | 0.002 |
| I would feel at ease around someone with cancer | 2.5(1.9) | 2.2(1.6) |  | 1.2(1.03) | 1.6(1.5) |  |
| I would feel comfortable around someone with cancer | 2.5(1.9) | 2.2(1.6) |  | 1.2(1.03) | 1.6(1.5) |  |
| I would find it difficult being around someone with cancer | 2.2(1.8) | 2.1(1.7) |  | 1.1(0.8) | 1.6(0.4) |  |
| I would find it hard to talk to someone with cancer | 1.9(1.5) | 1.8(1.4) |  | 1.2(1.06) | 1.6(0.5) |  |
| I would feel embarrassed discussing cancer with someone who had it | 2.2(1.7) | 2.1(1.7) |  | 1.3(1.1) | 1.9(1.8) |  |
| **Severity** | 4.2(1.4) | 4.05(1.5) | 0.306 | 1.7(1.2) | 2.9(1.7) | <0.001 |
| Once you've had cancer, you can never be 'normal' again | 4.03(1.2) | 3.8(1.8) |  | 1.6(1.4) | 3.06(2.1) |  |
| Getting cancer means having to mentally prepare oneself for death | 4.1(1.9) | 4.1(1.9) |  | 1.8(1.6) | 2.8(2.1) |  |
| Having cancer usually ruins a person's career | 4.7(1.6) | 4.4(1.8) |  | 1.8(1.6) | 3.2(2.2) |  |
| Cancer usually ruins close personal relationships | 4.08(1.9) | 3.9(2.07) |  | 1.7(1.5) | 2.4(2) |  |
| Cancer devastates the lives of those it touches | 4.1(1.9) | 3.9(2) |  | 1.7(1.5) | 3(2.2) |  |
| **Avoidance** | 1.9(1.1) | 2.02(1.1) | 0.813 | 1.1(0.6) | 1.5(1.1) | <0.001 |
| I would feel irritated by someone with cancer | 1.6(1.3) | 1.6(1.3) |  | 1.07(0.5) | 1.3(1.1) |  |
| I would avoid a person with cancer | 2.8(2.06) | 3(2.1) |  | 1.3(1.2) | 2.02(1.9) |  |
| I would distance myself physically from someone with cancer | 2.7(1.9) | 2.7(2) |  | 1.2(0.9) | 2.1(1.9) |  |
| If a colleague had cancer, I would try to avoid them | 1.2(0.9) | 1.4(1.09) |  | 1.1(0.6) | 1.1(0.7) |  |
| I would feel angered by someone with cancer | 1.3(1.09) | 1.2(0.8) |  | 1.05(0.4) | 1.2(1) |  |
| **Policy Opposition** | 1.2(0.4) | 1.3(0.5) | 0.289 | 1.03(0.2) | 1.05(0.2) | 0.405 |
| The needs of people with cancer should be given top priority | 1.1(0.3) | 1.2(0.5) |  | 1(0.09) | 1.04(0.2) |  |
| More government funding should be spent on the care and treatment of those with cancer | 1.1(0.6) | 1.2(0.5) |  | 1.02(0.2) | 1.04(0.2) |  |
| We have a responsibility to provide the best possible care for people with cancer | 1.3(0.8) | 1.5(1.1) |  | 1.05(0.4) | 1.07(0.4) |  |
| **Personal Responsibility** | 3.4(1.5) | 3.3(1.4) | 0.944 | 2.3(1.6) | 3(1.9) | 0.008 |
| A person with cancer is liable for their condition | 4.01(1.9) | 4.1(1.7) |  | 2.7(2.2) | 3.1(2.2) |  |
| A person with cancer is accountable for their condition | 3.9(1.8) | 4.1(1.7) |  | 2.8(2.2) | 3.1(2.2) |  |
| If a person has cancer, it's probably their fault | 2.8(1.9) | 2.6(1.8) |  | 1.9(1.7) | 2.8(2.1) |  |
| A person with cancer is to blame for their condition | 2.8(1.9) | 2.5(1.8) |  | 1.9(1.7) | 2.8(2.1) |  |
| **Financial discrimination** | 3.03(1.1) | 2.9(1.1) | 0.102 | 1.1(0.5) | 1.3(0.8) | 0.037 |
| It is acceptable for banks to refuse to make loans to people with cancer | 4.1(1.7) | 3.6(1.7) |  | 1.1(0.6) | 1.5(1.3) |  |
| Banks should be allowed to refuse mortgage applications for cancer-related reasons | 1.7(1.2) | 2(1.6) |  | 1.3(1.09) | 1.5(1.2) |  |
| It is acceptable for insurance companies to reconsider a policy if someone has cancer | 3.1(1.8) | 3.2(1.7) |  | 1.06(0.5) | 1.1(0.6) |  |
| **Overall stigma** | 2.7(0.7) | 2.6(0.7) |  | 1.4(0.6) | 1.9(0.9) | <0.001 |

<sup>a</sup> *p* values were calculated using Independent t-test
CASS scores range from 1 to 6; higher scores indicate greater stigma.

### Prevalence of cancer stigma

At baseline, the stigma prevalence was 26% in the intervention group and 28% in the control group. However, during follow-up, it decreased to approximately 4% in the intervention group and 13% in the control group. After the intervention, screening uptake was greater in the intervention group (82%) than in the control group (49%). Fig 2 shows the prevalence of cancer stigma, i.e., <3 (indicating no stigma) or ≥3 (indicating stigma).

**Fig 2.**
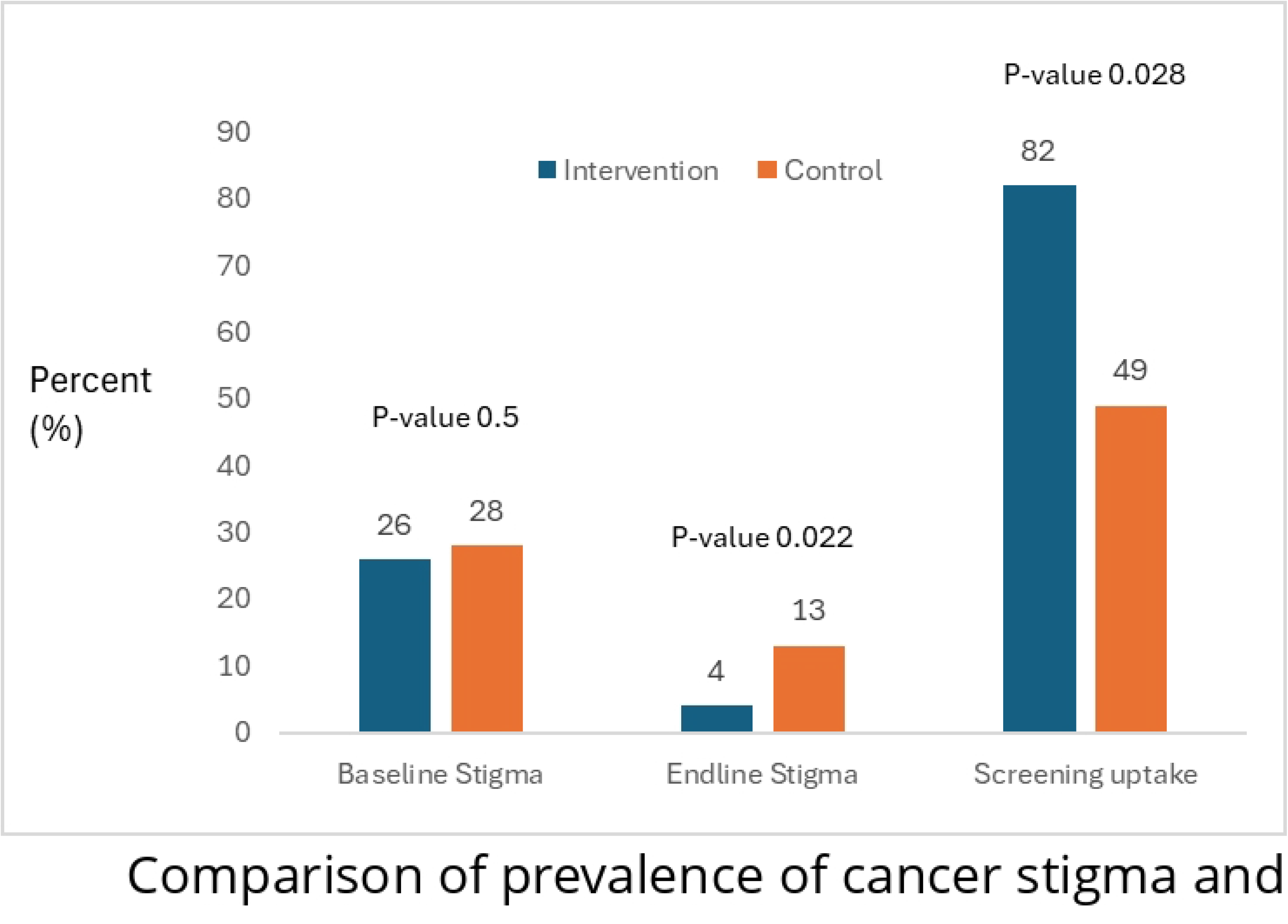
Comparison of prevalence of cancer stigma and screening uptake rates between intervention and control groups.

### Effect of the cervical cancer stigma reduction intervention on cancer stigma

In the intent-to-treat analysis, the odds of experiencing overall cancer stigma were 0.25 times lower in the intervention group than in the control group (95% CI: 0.08-0.8, p value = 0.022) after accounting for the clustering within wards. There was a significant reduction in cancer stigma in the two stigma domains. Specifically, the odds of cancer stigma were 0.21 times lower (95% CI: 0.08-0.56, p value = 0.001) in the awkwardness domain and 0.31 times lower (95% CI: 0.1-0.9, p value = 0.048) in the severity domain in the intervention group than in the control group. No significant differences were observed in the personal responsibility (p value = 0.549), avoidance (p value = 0.213), or financial discrimination (p value = 0.443) domains between the two groups. As treated analysis included 23 participants who were randomized to the intervention and declined. In this analysis, these participants were included in the control group.

The odds of cancer stigma were 0.09 times lower in the intervention group than in the control group (95% CI: 0.01-0.53, p value=0.007) after adjusting for age, education level, religion, ethnicity, work status, personal income, family income, and clustering within wards. Among the six cancer stigma subdomains, the intervention group had lower odds of stigma in terms of severity (OR=0.16, 95% CI: 0.06-0.42, p value < 0.001) and awkwardness (OR=0.12, 95% CI: 0.02-0.44, p value = 0.002). No significant differences were observed in the avoidance (p value = 0.141), personal responsibility (p value = 0.186), or financial discrimination (p value = 0.153) subdomains. At baseline, stigma in the policy opposition domain was reported by only 3 participants. By endline, none of the participants in either the intervention or control group reported stigma in this domain (Table 4)

**Table 4.** Effect of cervical cancer stigma reduction intervention on the cancer stigma.

| Stigma Domains | Intent to treat |  | As-treated |  |
| --- | --- | --- | --- | --- |
|  | OR (95% CI) | P value <sup>a</sup> | AOR (95% CI) | P value <sup>a</sup> |
| **Severity** |  |  |  |  |
| Control | Ref |  | Ref |  |
| Intervention | 0.31 (0.1-0.9) | 0.048 | 0.16 (0.06-0.42) | <0.001 |
| **Awkwardness** |  |  |  |  |
| Control | Ref |  | Ref |  |
| Intervention | 0.21 (0.08-0.56) | 0.001 | 0.12 (0.02-0.44) | 0.002 |
| **Personal Responsibility** |  |  |  |  |
| Control | Ref |  | Ref |  |
| Intervention | 0.74 (0.29-1.93) | 0.549 | 0.52 (0.19-1.4) | 0.186 |
| **Avoidance** |  |  |  |  |
| Control | Ref |  | Ref |  |
| Intervention | 0.44 (0.12-1.5) | 0.213 | 0.19 (0.02-1.7) | 0.141 |
| **Financial discrimination** |  |  |  |  |
| Control | Ref |  | Ref |  |
| Intervention | 0.54 (0.11-2.57) | 0.443 | 0.2 (0.02-1.79) | 0.153 |
| **Total stigma** |  |  |  |  |
| Control | Ref |  | Ref |  |
| Intervention | 0.25 (0.08-0.8) | 0.022 | 0.09 (0.01-0.53) | 0.007 |

<sup>a</sup> *p* values were calculated using Generalized Estimation Equation with logistic regression
\* For as-treated analysis adjusted for age (in years), level of education completed (formal and non-formal), religion (Hindu and others), ethnicity (Janjati, Brahmin/Chettri and others), work status (homemaker, job, daily wage labor, self-employed and others), personal income (in USD), and family income (in USD)

### Effect of the cervical cancer stigma reduction intervention on cervical cancer screening uptake

By six months, in the intent to treat analysis cervical cancer screening uptake was 3.9 times higher in the intervention group than in the control group (95% CI: 1.1-13.2, p value=0.028). The cervical cancer screening uptake was 4.02 times in the intervention group than in the control group (95% CI: 1.1–13.7, p value=0.026) in the as-treated analysis after accounting for clustering by ward and adjusting for age, education level, religion, ethnicity, work status, personal income, family income, age at pregnancy, number of pregnancies and history of menopause (Table 5)

**Table 5.** Effect of the cervical cancer stigma reduction intervention on cervical cancer screening uptake.

| Screening uptake | Intent to treat |  | As-treated |  |
| --- | --- | --- | --- | --- |
|  | OR (95% CI) | P-value | AOR (95% CI) | P-value |
| Control | Ref |  | Ref |  |
| Intervention | 3.9 (1.1-13.2) | 0.028 | 4.02 (1.1-13.7) | 0.026 |

*<sup>a</sup> p values were calculated using Generalized Estimation Equation with logistic regression*
*For as-treated analysis adjusted for age (in years), level of education completed (formal and non-formal), religion (Hindu and others), ethnicity (Janjati, Brahmin/Chettri and others), work status (homemaker, job, daily wage labor, self-employed and others), personal income (in USD), and family income (in USD), history of menopause, number of pregnancies, and age at first pregnancy.*

## Discussion

In this cluster-randomized controlled trial conducted in an urban municipality in Nepal, a community-based multicomponent intervention designed to reduce cancer stigma was associated with 0.25 times lower odds of having cancer stigma two months after the intervention and 4 times higher odds of cervical cancer screening during six months of follow-up compared with the control group. The results of the current study showed that a cervical cancer stigma reduction intervention was effective in reducing cancer stigma and increasing screening uptake among women aged 30-60 years in Nepal. This is particularly significant in Nepal, where screening coverage has remained stagnant for years[27].

The current study highlights the role stigma reduction interventions can play in addressing cancer stigma. Studies that have examined the impact of interventions targeting cancer stigma reduction have not thus far been available. We focused on parasocial relationships with cancer survivors, participatory discussions, and myth-busting activities. Similarly, stigma reduction approaches in the fields of mental health and HIV have employed survivor contact, education, and peer support with success[28]. When evaluating HIV-related stigma reduction interventions, participatory educational exercises seemed to reduce HIV misinformation stigma, although the effect weakened over a longer period [29]. Contact and educational interventions in the mental health field have led to small-to-medium reductions in stigmatizing attitudes[30]. Conversely, methods relying solely on education have proven ineffective in reducing cancer stigma, highlighting the complexity of stigma’s underlying drivers and the need for multicomponent interventions[31].A scoping review of the U.S. literature emphasized that addressing stigma drivers and fostering environments of self-efficacy and empowerment are key to improving screening rates[32]. Conversely, interventions focused only on education and brief health talks have shown no effect on increasing cervical cancer screening in Kenya and South Africa[17,33].

Our stigma reduction intervention was comprehensive and included multiple components such as; educational methods to enhance knowledge, a video featuring cervical cancer survivors was presented with the aim of fostering a parasocial relationship helping participants connect and relate to the life experiences of the survivors featured in the video and participatory group discussions to dispel misinformation and develop skills to address stigmatizing situations. These components together likely resulted in positive knowledge, attitudes, and behavior changes. Other studies have shown that focusing on knowledge alone does not necessarily increase screening rates or reduce stigma among women[18,29,30]. Educational interventions have been shown to be effective in reducing self-stigma but not public stigma[28]. Contact-based anti-stigma interventions, similar to our interventions, which focus on person-to-person interactions, have benefitted both the public and self-stigma by fostering empowerment and boosting self-esteem in a mental health context[28].

To our knowledge, this is the first study to assess the effectiveness of a cervical cancer stigma reduction intervention in reducing cancer stigma. The use of a standardized tool in Nepal, with a Cronbach’s alpha of 0.85, enhances the credibility of our findings. Inclusivity was ensured by facilitating the participation of women from diverse socioeconomic backgrounds across the study site. We used a generalized estimating equation model to address dependence of clusters in this study. Intent-to-treat and as-treated analyses were conducted, revealing reductions in stigma scores within the intervention group compared with the control group, further reinforcing the robustness of our findings.

This study has several limitations. First, blinding was not feasible. Second, the reach of intervention to the targeted participants was challenging, with almost 30% of participants declining the intervention due to festive seasons, elections, and difficulties in follow-up. Third, tracking participants was challenging as phone numbers were used as the major means of tracking which was unreachable and switched off most of the time. Fourth, there might be social desirability bias in reporting stigma. Finally, the long-term impact of the study is still unknown as follow-up occurred two months post intervention. Stigma reduction interventions have demonstrated weaker effects over extended periods[29]. Budanilkantha Municipality is an urban municipality in Nepal. Thus, the effectiveness of the intervention should be further tested in rural areas of Nepal.

## Conclusion

A community-based multicomponent intervention was associated with lower odds of elevated cancer stigma at 2 months and approximately fourfold higher odds of cervical cancer screening by 6 months among women in an urban municipality in Nepal. The findings provide preliminary evidence that stigma reduction may complement routine information-based approaches to cervical cancer prevention. Larger cluster randomised trials with longer follow-up are needed before widespread implementation.

## Ethics approval and consent to participate

Ethical approval was obtained from the Institutional Review Committee of Kathmandu University School of Medical Sciences (IRC reference 42/22). Written informed consent was obtained from literate participants. For participants who were illiterate, verbal informed consent was obtained and the consent form was signed by a witness chosen by the participant. Participants were informed of confidentiality, voluntary participation and their right to withdraw without consequences.

## Data availability

Dataset is publicly available at https://doi.org/10.6084/m9.figshare.33919306

## Funding

The screening program was supported by a grant “Machine Learning Based Cervical Screening Tool Development and Commercialization” funded by Kathmandu University Integrated Rural Development Program/ Nepal Technology Innovation Center (KU-IRDP/NTIC). All authors confirm the absence of any additional conflicts of interest.

## Competing Interests

The authors declare that they have no competing interests.

## Authors’ contributions

PT: Conceptualization, data curation, formal analysis, investigation and methodology project administration, resources, software, supervision, validation, visualization, writing-original draft, and writing-review & editing

BP: conceptualization, formal analysis, methodology, software, supervision, validation and writing-review & editing

MS: Data curation, investigation, project administration, resources and writing-original draft

AG: Data curation, investigation, project administration, resources and writing-original draft

SL: Formal analysis, methodology, software, validation, writing-original draft and writing-review & editing

LP: Investigation, project administration, resources, validation and writing-original draft

YSR: Conceptualization, formal analysis, methodology, supervision, validation writing-original draft and writing-review & editing

DS: Conceptualization, formal analysis, methodology, software, validation and writing-review & editing

SSS: Conceptualization, formal analysis, methodology, supervision, validation and writing-review & editing

AS: Conceptualization, formal analysis, methodology, software, supervision, validation and writing-original draft

ASh: Conceptualization, Data curation, Formal analysis, Investigation, Methodology, Project administration, Resources, Software, Supervision, Validation, Visualization, Writing-original draft and Writing-review & editing

## Acknowledgment

The authors thank the Department of Public Health and Community Programs, Kathmandu University School of Medical Sciences and the Budanilkantha Municipality public health team. We also thank ward chairpersons, elected female ward representatives and Female Community Health Volunteers for their support with community mobilization and screening activities.

## Supporting Information

S1 CONSORT Checklist. CONSORT checklist for the reported cluster-randomized trial.

S2 Ethics Approval. Approval letter from the Institutional Review Committee of Kathmandu University School of Medical Sciences.

S3 Study Protocol. Study protocol for the cluster-randomized trial.

S4 Data Collection Tools. Study questionnaires and data collection instruments.

S5 Consent Form. Participant informed consent documentation

## Notes

### Competing Interest Statement

The authors have declared no competing interest.

### Clinical Trial

ClinicalTrials.gov, NCT05489978

### Author Declarations

Ethical approval was obtained from the Institutional Review Committee of Kathmandu University School of Medical Sciences (IRC reference 42/22).

